# Cross-Linguistic Analysis of Speech Markers: Insights from English, Chinese, and Italian Speakers

**DOI:** 10.1101/2024.10.15.24314191

**Authors:** Gaia C. Santi, Eleonora Catricalà, Stephanie Kwan, Anson Wong, Zoe Ezzes, Lisa Wauters, Valentina Esposito, Francesca Conca, Daniela Gibbons, Eric Fernandez, Migual A. Santos-Santos, Chen TaFu, Kwan-Chen Li-Ying, R Lo, J Tsoh, Lung-Tat Chan, Adolfo M. Garcia, Jessica de Leon, Zachary Miller, Jet M.J. Vonk, Rose Bruffaerts, Stephanie M. Grasso, Isabel E. Allen, Stefano F. Cappa, Maria Luisa Gorno-Tempini, Boon Lead Tee

## Abstract

Cross-linguistic studies with healthy individuals are vital, as they can reveal typologically common and different patterns while providing tailored benchmarks for patient studies. Nevertheless, cross-linguistic differences in narrative speech production, particularly among speakers of languages belonging to distinct language families, have been inadequately investigated. Using a picture description task, we analyze cross-linguistic variations in connected speech production across three linguistically diverse groups of cognitively normal participants—English, Chinese (Mandarin and Cantonese), and Italian speakers. We extracted 28 linguistic features, encompassing phonological, lexico-semantic, morpho-syntactic, and discourse/pragmatic domains. We utilized a semi-automated approach with Computerized Language ANalysis (CLAN) to compare the frequency of production of various linguistic features across the three language groups. Our findings revealed distinct proportional differences in linguistic feature usage among English, Chinese, and Italian speakers. Specifically, we found a reduced production of prepositions, conjunctions, and pronouns, and increased adverb use in the Chinese-speakers compared to the other two languages. Furthermore, English participants produced a higher proportion of prepositions, while Italian speakers produced significantly more conjunctions and empty pauses than the other groups. These findings demonstrate that the frequency of specific linguistic phenomena varies across languages, even when using the same harmonized task. This underscores the critical need to develop linguistically tailored language assessment tools and to identify speech markers that are appropriate for aphasia patients across different languages.

## Introduction

There are over 7,000 living languages worldwide.^1^ Studying different languages can improve our knowledge of the neural underpinnings of speech and language functions,^2^ enhance our ability to diagnose and manage language symptoms in neurological diseases across diverse linguistic populations,^3^ and avoid health inequities stemming from the neglect of the impact of linguistic variations.^4^ While cross-linguistic studies have investigated different populations (i.e., cognitively normal adults,^5,6^ children across developmental stages,^7^ and participants with neurological diseases^3,8–13)^ across various tasks, most of these studies have only included languages of the same family (i.e., Indo-European; e.g., English vs German,^9^ Italian,^3,9^ or Spanish^11^). Even with crosslinguistic studies examine languages within the same language family, researchers have identified significant differences in the characterization of agrammatism across Indo-European languages, largely driven by morphological variations inherent to each language. Likewise, phonetic, and phonological differences significantly influence the characterization of motor speech impairments between English and Italian patients, particularly in relation to the complexity of word structure. Few studies have examined linguistic patterns across different language families (i.e., English vs Chinese,^14^ English vs Koean^13^) and have also identified notable differences in the frequency of produced tokens, such as pronouns or verbs, which contribute to the characterization of linguistic prototypical patterns. However, these studies are still scarce and would greatly benefit from comparing languages with more distinct typologies to better capture the full spectrum of linguistic diversity. To overcome this existing knowledge gap and establish benchmarks useful for developing linguistically tailored diagnostic tools, this study examined the connected speech production of cognitively normal speakers of English, Chinese (Cantonese and Mandarin), and Italian languages from different families with varying degrees of linguistic proximity.

English and Italian stem from distinct branches of the Indo-European family — Germanic and Roman, respectively, while the Chinese languages belong to the Sino-Tibetan language family.^15,16^ According to eLinguistic (2023), which quantifies the temporal distance of when two languages last shared a common ancestor (i.e., language genetic proximity), English and Italian languages have a high language-relatedness. In contrast, English and Chinese, as well as Italian and Chinese, do not show a shared linguistic ancestry. These language distance reflect the degree of linguistic similarities across various linguistic domains, including but not limited to phonology, morphology and syntax.^17^ For instance, in terms of phonology, the English language is organized into consonant-vowel (CV) or CCV clusters; Italian predominantly utilizes a CV cluster, and Chinese relies on V, CV, or CVC configurations.^18–20^ Furthermore, Chinese is a tonal language requiring the control of oropharyngeal and vocal cord muscles to generate pitch variations that synchronize with syllable production. In contrast, English and Italian rely on lexical stress, emphasizing specific syllables through increased voice intensity, vowel length, and changes in pitch. At the level of word morphology, Italian has richer inflectional morphology than English,^21^ while Chinese is an analytic language lacking affixes and having only very few types of inflectional and derivational morphology. Moreover, unlike English and Italian, the Chinese language requires the use of mandatory classifiers when numbers or determiners precede nouns.^22,23^ Syntactic differences, although understudied,^24^ are also present. For example, Italian and Chinese are pro-drop languages: elements such as pronouns, verbs, and/or determiners are not always obligatory to form grammatically coherent sentences.^25,26^ Variations across these and other linguistic features can significantly impact speech production and, consequently, its assessment.

In this study, we aim to profile the intrinsic characteristics of each language and to identify linguistic differences and similarities across these three languages in response to a single, harmonized task, such as the widely used picture description task. This task is commonly used in both clinical and research settings across cultures, yet normative cross-linguistic data remain absent. Connected speech production provides insights across various linguistic aspects, including phonological, lexico-semantic, morpho-syntactic, and discourse/pragmatic domains.^27^ This knowledge can be instrumental in unraveling the role of linguistic diversity and potentially contribute to the development of accurate language assessments, particularly in cases of neurological impairment.^4^ The present study had two primary objectives: 1) to delineate the distinctive linguistic characteristics inherent to English, Chinese, and Italian speakers, and 2) to identify shared linguistic features among these languages, with the goal of developing language assessment tools that have cross-linguistic applicability, particularly for assessing patients with aphasic production, such as those with neurodegenerative conditions.

## Materials and Methods

### Participants

A cohort of 39 cognitively normal (CN) participants was included in the study, with 13 individuals representing each of the target languages. Older adults (aged 50 years and above) were selected as they provide an ideal benchmark for comparison with populations affected by aphasia and other neurological diseases. The cohort included American English speakers from the University of California, San Francisco, Chinese speakers from the Chinese Language Assessment Project (CLAP) collected across seven sites in Taiwan and Hong Kong,^23^ and Italian speakers through collaboration with the IUSS Cognitive Neuroscience (ICoN) Center at the Institute for Advanced Studies, IUSS Pavia. All included CN participants had no history of neurological or psychiatric disorders and were native speakers of one of the included languages. Each participant completed an oral picture description task, specifically the oral description of the picnic scene from the Western Aphasia Battery.^28^

### Oral picture description

The picture description task is a widely used tool among neuropsychologists and speech-language pathologists due to its time efficiency and minimal technical requirements, while providing rich language data across various linguistic aspects (motor speech, phonology, lexical semantics, and morpho-syntax). Additionally, using a picture scene to prompt oral production helps access conceptual knowledge in both cognitively normal individuals and those with neurological diseases.^29,30^ Performance data were audio recorded and securely stored in accordance with the data protection protocols of each participating center. The examiners instructed the participants to observe the picture and describe what they saw in sentences. If the participant paused before one minute of production, the examiner prompted them to continue by asking if he/she could describe more of what they observed, using sentences. The audio samples were then manually transcribed, and linguistic features of interest (as detailed in Section 2.3) were coded according to the Computerized Language ANalysis program (CLAN) instructions, as outlined in Section 2.4. Two raters independently transcribed and coded the speech samples for each language group. The results were then compared, and any coding discrepancies were addressed by consulting a third rater. The raters discussed each case individually until a consensus was reached.

### Linguistic features

A total of 28 features was chosen (listed in Table 1 with definitions), drawing upon existing literature^3,31–33^ and the authors’ native knowledge of each language (American English: ZE, LW; Chinese SW, AW, BL; Italian GCS, VE, EC). These features encompass phonology, lexico-semantic, morpho-syntactic, and discourse and pragmatics. For each domain, we selected features that were considered relevant for at least one of the languages included in the study, in particular they were: 1) features that have been previously used to describe the linguistic performance of healthy participants;^33^ 2) features that characterized the linguistic performance of clinical populations, i.e., stroke aphasia^12^ and primary progressive aphasia patients.^27,32^

**Table 1.** Speech and Language included features.

| Domains | Features | Definition |
| --- | --- | --- |
| Phonological | N° word repetitions/tot words* | Starts to say a word, stop, and repeat without changes |
| Data | N° prolonged sounds* | Sounds that are abnormally prolonged |
|  | N° broken words (or within word pause) * | Pause within word |
|  | N° abandoned words/tot words* | Starts to say one word but then says a different word instead |
|  | N° empty pauses/tot words* | Number of silent (no sound produced) pauses |
|  | N° filled pauses/tot words* | Number of filled pauses (i.e., uhm, ehm) |
| Lexico-semantic | - open class words/total words | Total number of content words (verbs, nouns, adjectives, adverbs)/total number of words |
|  | - closed class words/total words | Total number of function words (determiners, classifiers<sup>§</sup>, pronouns, prepositions...)/total number of words |
| Morpho-syntactic | N° utterances<sup>†</sup> | Utterances require implicit/explicit subject and main verb |
|  | Mean Length of Utterances | Ratio of morphemes to utterances |
|  | N° utterances without verb/N° utterances *<sup>°</sup> | Elision (drop) of main verb when not grammatically permitted |
|  | Determiner elision<sup>°</sup>/substitution* | Elision (drop) or substitution of article |
|  | Prepositions elision<sup>°</sup>/substitution* | Elision (drop) or substitution of preposition |
|  | Classifier elision<sup>§°</sup>/generalization<sup>§°</sup> | Elision (drop) or generalization of classifier |
| Discourse and pragmatic | speech rate* | Total number of words/minutes |
|  | N° irrelevant words/tot words* | N° words syntactically correct but do not convey relevant meaning/ total number of words |
|  | N° tangential words/tot words* | N° words not related to the task/total number of words |

List of include features and their definition.
\* Features manually coded.
<sup>§</sup> specific features for Chinese language.
<sup>°</sup> counted only if it is an error for the respective language
<sup>†</sup> Adapted for Chinese and Italian
N° number.

Three out of the 28 features – namely, the ratio of classifiers over the total number of words, the number of classifier generalizations, and the number of classifier omissions – were specific only to Chinese speakers. Additionally, the definition of utterance taken in to account linguistic diversity, while three other features – the number of utterances without verbs, the number of agrammatic determiner elision, and preposition elision – were counted only in case they represented an error for the respective language, as follows:

− Utterances are defined, according to CLAN manual, as sentence with subject and main verb. The main clause together with dependent clauses, adjuncts, and adverbial phrases were counted as one utterance. Both Italian and Chinese allow the drop of the subject. Therefore, in these two languages, utterances were counted and considered correct if the main verb was present. For example, the following “She is reading a book” is counted as one utterance in English. In Italian, instead, one could count as utterances “ Sta leggendo un libro,” or in Chinese, “在看書” instead of “她在看書.”
− In English, nominal sentences (i.e., a sentence without a finite verb) are relatively uncommon. Conversely, both Italian and Chinese sentences, can be considered more frequently as grammatically correct even without a verb. In Italian, the main verb can be implied if repeated across coordinate/subordinate sentences, as in the example “Vedo una coppia. Un albero con tante foglie” (literal English translation: I see a couple. (I see) A tree with many leaves). In Chinese, adjectival sentences can be formed without a verb, as shown in “這棵樹很高” (literal English translation: “This [classifier] tree very high”). While English requires a verb, as in “This tree <u>is</u> very high,” where the verb “is” is essential, the mandatory classifier use in Chinese is not necessary in English.
− In both English and Italian, determiners are frequently used to convey specificity, definiteness, and other nuances in meaning, making them an integral part of sentence construction. Consequently, the omission of determiners often results in ungrammatical sentences. However, in Chinese, determiners are less commonly used, and their omission can often still yield grammatically correct sentences. For example, in Chinese, “她買了車” (literal English translation: “She bought car;” literal Italian translation: “Ha comprato macchina”) remains grammatically intact even without a determiner “a” or “una” before the word “車/car/macchina.”
− The omission of prepositions often results in ungrammatical sentences in English and Italian, whereas in Chinese, the grammatical structure can still remain intact without them. For instance, in English, you would say “She is going to the store,” and in Italian, “Sta andando al negozio.” However, in Chinese, it is acceptable to say “她去商店” (literal English translation: “She is going store”) without the preposition “到.” Unlike in English and Italian prepositions (e.g., “to” and “al”), the preposition can be omitted in Chinese without compromising grammaticality.

### Computerized Language ANalysis program

Speech samples were manually transcribed and coded using CLAN, following the instructions provided in its manual. CLAN, designed specifically for analyzing linguistic data, has been widely used in studies involving cognitively normal participants, children, and individuals with neurological diseases.^34^ For detailed information on CLAN and its applications, please refer to the Aphasia Bank project at https://aphasia.talkbank.org/. The CLAN software is available in multiple languages, including English, Chinese, and Italian, allowing us to work within a unified analytical framework. Speech production analysis was conducted using the CLAN pipeline, which enabled the automatic extraction of the features of interest. Features not pre-configured in the CLAN software were manually coded and then processed via the CLAN command interface. It is important to note that definitions and coding procedures may vary across the studied languages. An ad hoc code was used to classify certain features as errors in one language, while acknowledging that these same features might be acceptable in others (see the list in the linguistic features section and Table 1).

### Statistical analysis

Descriptive statistics for demographic data and linguistic features were calculated and compared across groups using chi-squared tests for categorical variables and ANOVA for continuous variables, with Bonferroni’s correction applied for post-hoc analyses. Given the specificity of some markers in characterizing the linguistic profile of one group, i.e., n° empty pauses/total words, we performed exploratory correlation analysis between distinctive features for each language (see supplementary materials).

## Results

### Demographic data

Demographic information for the healthy participants is presented in Table 2. Sex, age, and education levels were matched across the three language groups (English, Chinese, and Italian), with all p-values exceeding 0.05, indicating no significant differences.

**Table 2.** Demographic Data of healthy participants.

|  | English (n=13) | Chinese (n=13) | Italian (n=13) | P value |
| --- | --- | --- | --- | --- |
| Age (years) | 70.5 (3.2) | 67.53 (5.9) | 64.9 (8.2) | 0.080 |
| Education (years) | 16.6 (1.5) | 15.07 (1.6) | 14.7 (3.0) | 0.068 |
| Sex (Female/Male) | 4/9 | 4/9 | 5/8 | 0.749 |

*Mean and standard deviation, between brackets, are reported for each group or frequency/total.*

### Speech sample analysis: Language features

The results of the speech sample analysis across groups are summarized in Table 3. Among the 28 features examined, there were six features found to be significantly different across the three groups after Bonferroni’s correction (see Figure 1). Within the phonological domain, Italian speakers produced a higher ratio of empty pauses to total words than their English and Chinese counterparts (F(2,28)=11.98, p<0.001). In the lexico-semantic domain, the Chinese group produced a lower proportion of prepositions, determiners, and pronouns to total words compared to both English and Italian groups (F(2,28)=34.32, p<0.001; F(2,28)=307.26, p<0001; F(2,38)=16.508, p<0.001, respectively). The Italian group produced a lower proportion of prepositions compared to English speakers (p<0.001). Italian speakers generated a higher proportion of conjunctions than the English and Chinese groups (F(2,28)=18.31, p<0.001). Furthermore, the Chinese group used a higher proportion of adverbs compared to the English and Italian groups (F(2,28)=12.491, p<0.001).

**Figure 1.**
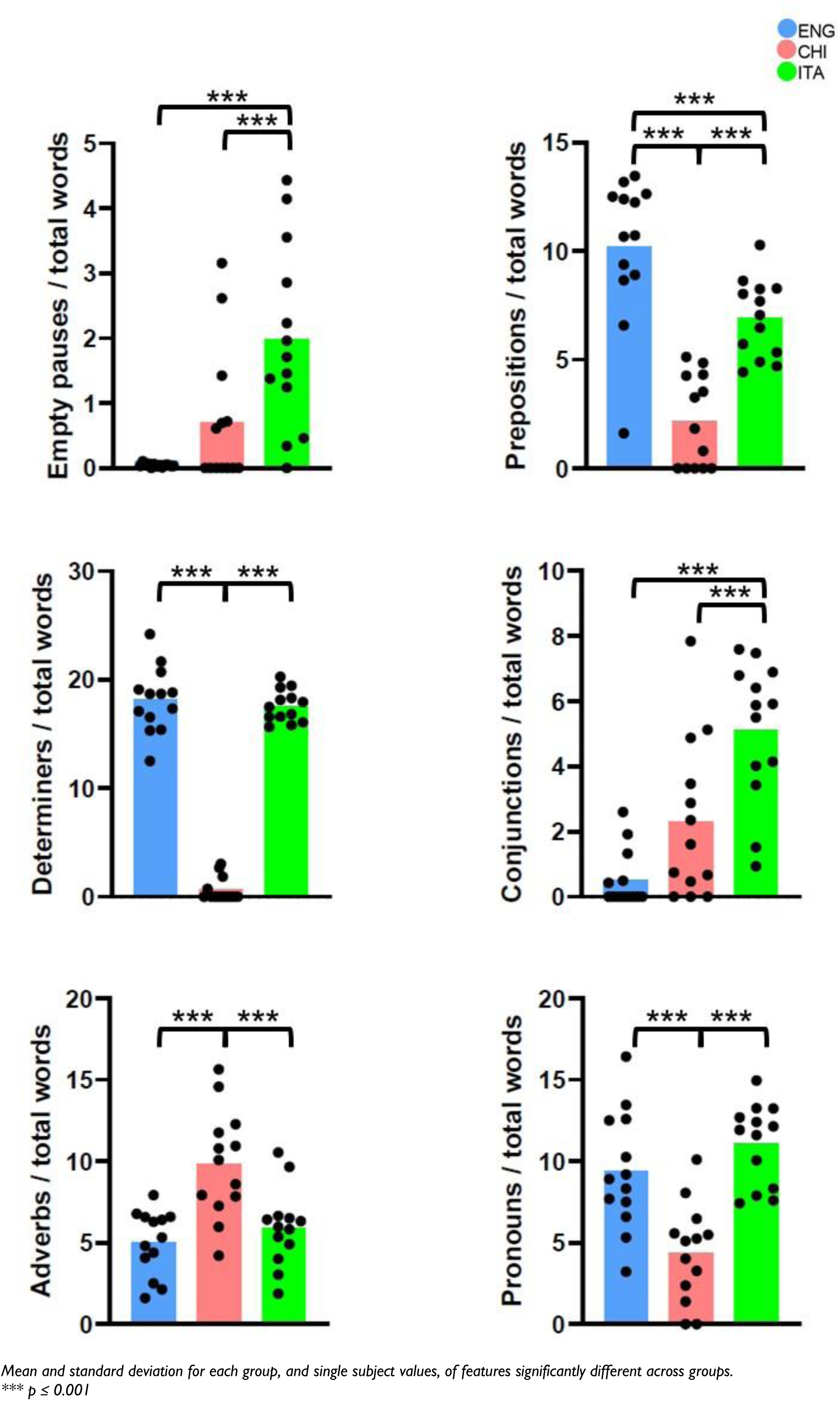
Scatter dot plot of significantly different features across languages. *Mean and standard deviation for each group, and single subject values, of features significantly different across groups. *** p ≤ 0.001*

**Table 3.**
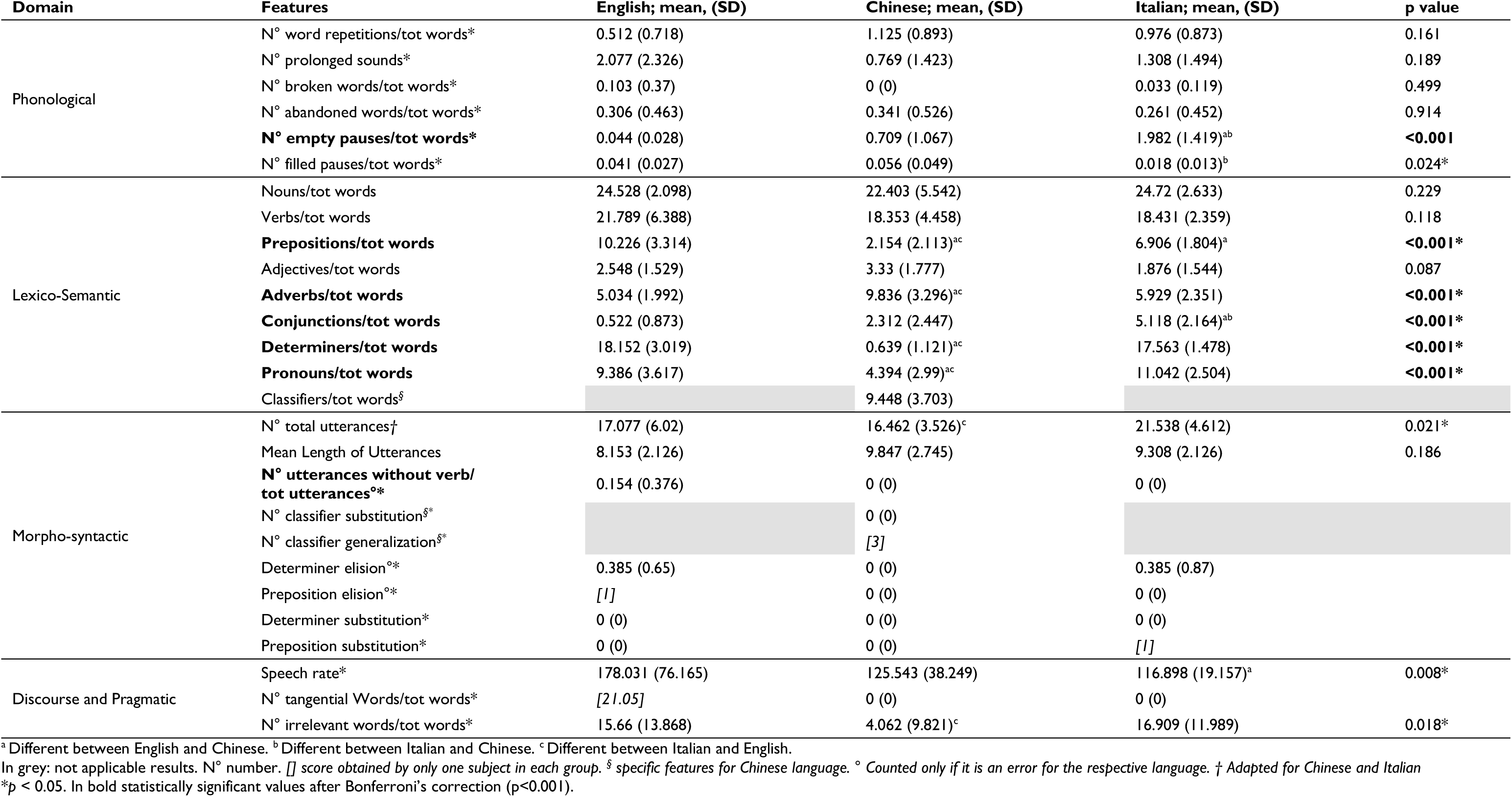
Crosslinguistic comparison of speech and language features across English, Chinese and Italian speakers.

In addition, several linguistic characteristics were found to be exclusive to certain languages. Specifically, the high proportion of classifiers usage and their generalization were uniquely present in Chinese speakers. Additionally, only English, and Italian cognitively normal speakers were noted to produce broken words. Due to the pro-drop nature of Italian and Chinese languages, verb-less utterances were acceptable in these languages and only considered incorrect when produced by English speakers. Finally, the remaining nineteen features, spanning various linguistic domains, were universally expressed across the three languages, with no significant differences after post-hoc multiple comparison correction.

To assess the individual heterogeneity within each group, we also examined unique deviations from the typical patterns observed within their respective language groups. Notably, only one participant in the English group exhibited preposition elision, and only one Italian speaker demonstrated preposition substitution. Additionally, only one of the Chinese speakers was noted for producing three instances of noun classifier generalizations—a feature not observed in any other subjects within that group. At the discourse level, only one English participant displayed tangential speech, constructing grammatically correct sentences that, nonetheless, did not relate to the content depicted in the picture, such as, “It looks like a small lake in Connecticut where we used to go.”

## Discussion

This study compared connected speech production of English, Chinese, and Italian healthy speakers using the widely utilized picture description task, a tool for which cross-linguistic norms are still lacking. Most existing cross-linguistic studies involve languages belonging to the same family, thus offering a limited perspective due to the inherent similarities among the languages.^3,35^ Few studies included a direct comparison of languages belonging to different families, such as English vs Chinese^12^ or English vs Korean.^13^ Results suggest that the verbal descriptions of the same pictures by cognitively normal individuals show some language-specific features. Yiu et al. found that the production of elliptical sentences was more frequent among healthy Chinese speakers. Sung et al. noted that Korean-speaking aphasic patients produced more verbs compared to English aphasic participants. Our paper represents an opportunity to increase the linguistic diversity, particularly through the inclusion of morphologically rich and limited languages such as Italian and Chinese, respectively. The consideration of cross-linguistic variation is crucial for the assessment of the manifestations of language impairment in clinical conditions, such as aphasias due to stroke or neurodegenerative diseases.

Our primary objective was to identify both shared and distinctive linguistic features that pertain to each language during the picture description task, thus offering valuable insights into the cross-linguistic applicability of this widely used tool. Our results underline several cross-linguistic variations and similarities that manifest in cognitively normal individuals matched for age, education, and sex. The primary differences were observed in the phonological and lexico-semantic domains, likely stemming from the inherent linguistic characteristics of each studied language. For a summary, please refer to Table 4. These findings hold significant importance in the development of speech markers aimed at identifying various neurological diseases across diverse linguistic contexts.

**Table 4.**
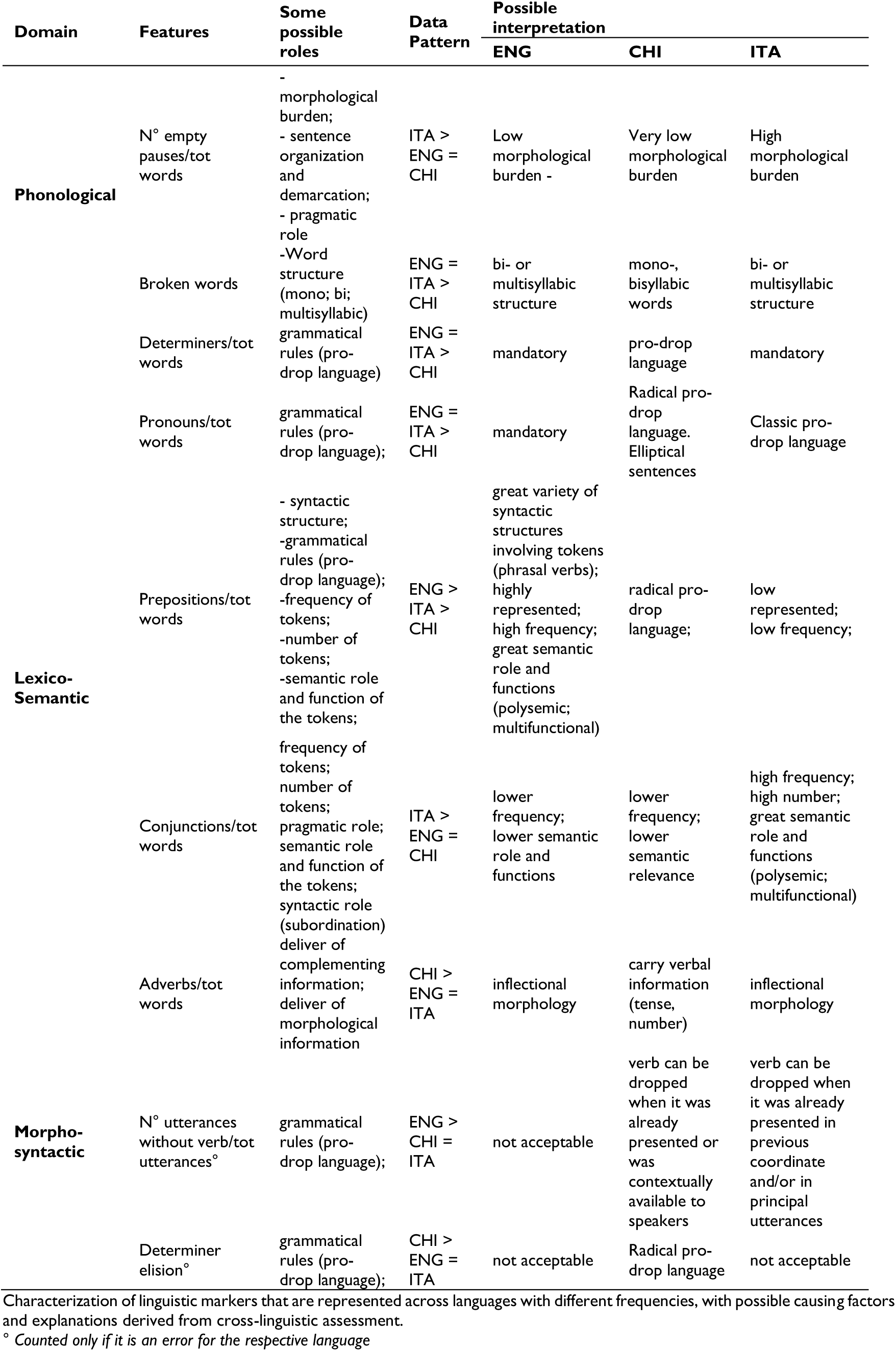
Cross-linguistic variabilities across English, Chinese, and Italian languages.

First of all, our results revealed significant differences in the phonological domain between the three groups. In particular, participants exhibited distinct patterns of pausing related to word segmentation: both English and Italian speakers displayed a similar proportion of pauses within words, resulting in “broken words,” whereas Chinese speakers exhibited none. This difference may be attributed to fundamental variations in word structure. Indo-European languages, such as English and Italian, often include bi- or multisyllabic words, while Chinese primarily relies on monosyllabic characters or disyllabic compound words.^18^ The multisyllabic nature of the Indo-European languages likely increase the complexity of phonological processing, potentially reducing fluency. An increase number of pauses in connected speech has been identified as an important diagnostic feature and early marker in vascular and/or neurodegenerative aphasias.^36–38^ These findings underscore the importance of a tailored cross-linguistic assessment of patient performance, as such features may not be similarly represented across different languages.

In addition to cross-linguistic differences between language families, we also identified variations in the phonological domain within the two Indo-European languages. Our results indicate that Italian speakers produce a higher proportion of empty pauses compared to both English and Chinese speakers. Empty pauses in speech production may serve numerous roles. A “strategic” pause may help in preparing the articulation of sounds, marking the demarcation of discourse, gathering thoughts, and structuring verbal expression, particularly when complex morphosyntactic structures are involved.^39,40^ Morphological complexity may be particularly relevant in interpreting these results, as it may create different demands for pauses and potentially explain the observed differences across language groups (see also Supplementary Material, Table 1). Studies on healthy speakers reported a relationship between disfluency and cognitive load.^41^ Research in aphasic patients supports similar conclusions, suggesting that increased pausing time is associated with deficits in sentence and language planning.^36^ Accordingly, the production of morphologically inflected words presents a greater phonological challenge compared to non-inflected words or words relying on derivational morphology.^42^ Italian, with its complex morphological structure,^21^ stands in stark contrast not only to Chinese—an analytic language—but also to English, which features more limited inflectional morphology.^43^

Furthermore, we identified numerous cross-linguistic differences in the lexical-semantic domain, particularly regarding the proportion and types of tokens predominantly used by speakers. This variation encompasses both functional words (e.g., determiners, pronouns, prepositions, and conjunctions), and content words (e.g., adverbs). Specifically, English, and Italian speakers tend to use more determiners, pronouns, and prepositions, whereas Chinese speakers produce a higher number of adverbs. Additionally, when comparing the two Indo-European languages, English speakers utilized more prepositions, whereas Italian speakers favored conjunctions.

Cross-linguistic variations in morphology, grammar and syntax may explain the differing usage patterns of tokens across languages. The possibility of omitting specific tokens (e.g., determiners, pronouns, conjunctions, and prepositions) varies by language and we speculate this variation plays a key role in interpretating our results. Chinese, a radical pro-drop language,^44^ commonly omits determiners and pronouns without compromising its grammaticality, unlike in English or Italian, where such omissions would often render sentences agrammatic.^45^ This phenomenon is reflected in the prevalent use of elliptical sentences in Chinese,^26,46–48^ suggesting that while pronouns or determiners are important, their usage may be comparatively lower than in English and Italian.^49^ This is also indirectly supported by studies on second language acquisition which have highlighted the complexity posed by determiners use,^50,51^ suggesting that Chinese speakers often erroneously omit determiners when speaking English.^52^ Similar phenomenon can also be extended to the disparate use of adverbs across the studied languages. We found that Chinese speakers employed adverbs more frequently than English and Italian speakers. This finding aligns with the characteristics of Chinese morphology. Unlike Italian and English, which use verb tenses, plural and gender forms of nouns (the latter only in Italian), Chinese does not convey chronological and numerical information through inflectional morphology or suffixes. Instead, this information is typically indicated by adverbs or aspect markers.^53^ These findings indicate the need for careful consideration of the diagnostic features of agrammatism in speakers of Chinese languages. The omission of “function words,” traditionally a defining characteristic of agrammatism,^54^ must be reevaluated in light of the unique morphosyntactic features specific to the language.^10^

Moreover, Italian speakers used a higher number of conjunctions compared to other studied languages. Indo-European languages such as Italian, typically rely on conjunctions to maintain cohesion, whereas Chinese languages achieve discourse coherence primarily through context and repetition.^46,55^ For instance, the English sentence “She went to the store and bought milk” can be translated into Chinese as “她去商店買牛奶,” where “並” (and) could be omitted, as the relationship between clauses is often inferred from the context. Consequently, native Chinese speakers were noted to more frequently omit conjunctions preceding subordinate clauses when speaking English, a feature that is grammatically inaccurate in English and uncommon among native English speakers.^56^ In Italian, conjunctions serve multiple grammatical and pragmatic functions, supporting both sentence structure and discourse organization.^57^ As a result, translation studies have shown that Italian translators rely more on conjunctions when translating into English to maintain speech coherence.^58^ Furthermore, many Italian adverbs can also function as conjunctions, hence significantly increasing their usage, particularly in spoken language.^57,59^

On the other hand, prepositions play a pivotal role in the English language, where approximately one in every ten words in English is a preposition,^60^ with a total of more than 100 prepositions - a number higher than in other languages.^61^ In contrast, Italian features only nine simple prepositions, and Chinese comprises twenty-five.^62^ Given the disparity in the number and frequency of prepositions across the studied languages, it is unsurprising that English speakers exhibited a notably greater use of preposition in our study. These token differences must be carefully considered when defining diagnostic features of language impairment across various languages.^63^

While numerous language-specific features were noted in our study, it is also important to emphasize that we identified a set of features across various linguistic domains that were uniformly distributed among all groups. In the phonological domain, the frequency of abandoned words, repeated words, and filled pauses exhibited no significant differences between the groups. Similarly, in the lexico-semantic domain, the distribution of nouns, verbs, and adjectives were consistent across all groups. In the morpho-syntactic domain, after adjusting for multiple comparisons, there were no significant disparities in the number of utterances or their lengths. At the discourse level, none of the studied characteristics showed significant variations between groups. These consistent findings suggest that these features may function as universal markers and should be considered in the cross-linguistic assessment of language disorders.

We acknowledge the limitations posed by the small sample size in our study, a common challenge in cross-linguistic research due to the need to match participants across various languages. Furthermore, our analysis incorporated a broad set of features to examine multiple linguistic domains, which might have affected the statistical power. In addition, we believe the features build on one another and reinforce each other when statistically significant. To limit the possible errors derived from this approach and ensure higher reliability of data, we adopted a stringent control method, using the Bonferroni’s correction. We also identified features that reflect individual heterogeneity within each language group, exemplified by classifier generalization among Chinese speakers, preposition omission or substitution, and tangential speech among English and Italian speakers. This variability may stem from the small sample size, the infrequent occurrence of certain linguistic phenomena, and diverse personal experiences, including differences in educational attainment and other individual factors. Therefore, sociodemographic factors must be carefully considered when analyzing speech and language markers.

Future studies should aim to expand the number of languages studied. Based on our findings, we believe that speech and language assessment tools or strategies should be developed with adequate consideration to linguistic typology when applied across different languages. The modality to elicit connected speech also warrants careful consideration. For instance, describing a complex picture, such as the “picnic picture” used in our study, is a common clinical assessment tool, but its cultural relevance may vary across different linguistic groups.^64^

In conclusion, we identified a set of linguistic markers that are distinctive across the three studied languages, alongside identifying shared linguistic features that may serve as potential universal markers for speech and language elicited by a picture description task. Taken together, our findings affirm that cross-linguistic differences are discernible even among cognitively normal participants when describing the same picture. Comparative studies across languages are crucial as they reveal essential typological patterns and provide customized benchmarks vital for disease diagnosis. This underscores the importance of adopting a cross-linguistic framework to ensure that speech and language assessments are effectively tailored to address unique linguistic symptoms and support the development of universally applicable cross-linguistic diagnostic tools.

### Data availability

Data are available under reasonable request to the corresponding author.

## Acknowledgments

We thank all the participants for their support and availability to complete the assessment.

## Participants’ consent

Participants consent was obtained according to the Declaration of Helsinki and was approved by the ethical committee of the institution in which the work was performed. Namely: Chinese University of Hong Kong, Ref No.: 2019.499, Title: Establishing evidence to speech and language intervention of Cantonese Speakers with Primary Progressive Aphasia (PPA); The Education University of Hong Kong, Ref No.: 2023-24-0338, Title: Chinese Language Assessment in Primary Progressive Aphasia: Cantonese (CLAP-C); National Taiwan University Hospital, NTUH-REC No. 01812096RINA, Title: Chinese Language Assessment in Neurodegenerative disease; Hualien Tzu Chi Hospital, REC No.: IRB111-222-B, Title: Chinese Language Assessment in Neurodegenerative disease; University of California San Francisco, No.: 10-03946, Title: Frontotemporal Dementia: Genes, Images and Emotions; IRCCS Mondino Foundation, Dementia Research Center, N. 20200026675; Title: Paradigmi Comportamentali per lo studio delle funzioni linguistiche e di memoria semantica nelle patologie neurodegenerative.

