## Supplementary Material for "Cross-Linguistic Analysis of Speech Markers: Insights from English, Chinese, and Italian Speakers"

SUPPLEMENTARY MATERIALS

### Feature Coding

N° utterances without verb/n° utterances: was calculated manually using data derived from CLAN analysis. Namely, we computed the number of senteces in which 0v (no-verb) code was present over the total number of utterances.

Classifier elision: was reported in CLAN using 0cl code.

Classifier generalization: was reported in CLAN using [* c] code.

### Correlation

We performed a correlation analysis between distinctive features, namely n° empty pauses/total words, derived from the speech production of healthy speakers of each language, namely English, Chinese, and Italian.

**Table 1 Correlation analysis**

| | **English** | **Chinese** | **Italian** |
|---|---|---|---|
| **Features** | **N° empty pauses/ tot words** | | |
| N° word repetition/tot words | -0.311 | -0.340 | 0.439 |
| N° prolonged sounds | -0.244 | -0.178 | -0.091 |
| N° broken words/tot words | 0.251 | | -0.058 |
| N° abandoned words/tot words | -0.500 | -0.294 | 0.089 |
| N° filled pauses/tot words | 0.220 | 0.127 | 0.172 |
| Nouns/tot words | 0.272 | -0.288 | 0.488 |
| Verbs/tot words | -0.046 | -0.191 | -0.263 |
| Preposition/tot words | 0.248 | 0.087 | -0.322 |
| Adjectives/tot words | -0.336 | -0.182 | 0.249 |
| Adverbs/tot words | 0.213 | -0.282 | -0.178 |
| Conjunctions/tot words | 0.515 | 0.234 | 0.071 |
| Determiners/tot words | -0.335 | -0.089 | **0.671** |
| Pronouns/tot words | -0.191 | -0.078 | -0.548 |
| Classifiers/tot words | | 0.074 | |
| N° total utterances | 0.081 | -0.053 | 0.475 |
| Mean Length of utterances | -0.415 | -0.303 | **-0.618** |
| Words/min | 0.272 | -0.396 | -0.203 |
| N° words empty speech/tot words | -0.197 | -0.202 | -0.155 |

In bold significant results ($p<0.05$)
In grey: not applicable results.
